# A multi-modal deep learning framework for predicting PSA progression-free survival in metastatic prostate cancer using PSMA PET/CT imaging

**DOI:** 10.1101/2025.07.12.25331141

**Authors:** Hamid Ghaderi, Chenyang Shen, Wadih Issa, Martin G. Pomper, Orhan K. Oz, Tian Zhang, Jing Wang, Daniel X. Yang

## Abstract

PSMA PET/CT imaging has been increasingly utilized in the management of patients with metastatic prostate cancer (mPCa). Imaging biomarkers derived from PSMA PET may provide improved prognostication and prediction of treatment response for mPCa patients. This study investigates a novel deep learning-derived imaging biomarker framework for outcome prediction using multi-modal PSMA PET/CT and clinical features. A single institution cohort of 99 mPCa patients with 396 lesions was evaluated. Imaging features were extracted from cropped lesion areas and combined with clinical variables including body mass index, ECOG performance status, prostate specific antigen (PSA) level, Gleason score, and treatments received. The PSA progression-free survival (PFS) model was trained using a ResNet architecture with a Cox proportional hazards loss function using five-fold cross-validation. Performance was assessed using concordance index (C-index) and Kaplan-Meier survival analysis. Among evaluated model architectures, the ResNet-18 backbone offered the best performance. The multi-modal deep learning framework achieved a 5-fold cross-validation C-index ranging from 0.75 to 0.94, outperforming models incorporating imaging only (0.70–0.89) and clinical features only (0.53–0.65). Kaplan-Meir survival analysis performed on the deep learning-derived predictions demonstrated clear risk stratification, with a median PSA progression free survival (PFS) of 19.7 months in the high-risk group and 26 months in the low-risk group (*P* < 0.001). Deep learning-derived imaging biomarker based on PSMA PET/CT can effectively predict PSA PFS for mPCa patients. Further clinical validation in prospective cohorts is warranted.

## Introduction

Prostate cancer is one of the leading causes of cancer-related deaths among men in the United States and worldwide.[1–3] Metastatic prostate cancer (mPCa) can have a heterogeneous disease course, with substantial variations in patient outcomes that are not fully captured by current clinical classification systems such as disease burden. While advancements in systemic and local therapies have significantly improved patient outcomes, the diverse disease trajectories and evolving treatment resistance of mPCa make prognostication, personalized treatment selection and therapeutic monitoring challenging.[4–7]

Imaging biomarkers have demonstrated considerable potential in predicting clinical outcomes and optimizing treatment strategies by providing non-invasive insights into disease progression and treatment response.[8–10] In recent years, prostate-specific membrane antigen (PSMA)-targeted positron emission tomography (PET/CT) imaging has emerged as an diagnostic imaging modality that has rapidly supplanted conventional imaging techniques due to its increased sensitivity and specificity.[11–13] It is increasingly recognized as an advanced imaging tool offering improved detection of metastatic lesions. However, its full prognostic value and the extent to which PSMA PET findings can predict treatment response and disease progression are still being explored. Developing robust tools for further risk stratification and interpretation of PSMA PET findings will be crucial for personalizing treatment strategies for mPCa patients.

There is considerable interest in utilizing artificial intelligence (AI) to derive PSMA PET-based imaging biomarkers in mPCa. Earlier approaches have utilized radiomics-based machine learning models.[9,14–19] Recent advances in deep learning have enhanced AI-driven outcome prediction, with researchers using deep neural networks to extract complex imaging patterns and for downstream patient outcome predictions. A previous study by Zhao et al. employed a 3D DenseNet convolutional neural network architecture for lesion-level malignancy classification and survival prediction from [^18^F]DCFPyL PSMA PET/CT imaging.[20] Others have applied neural networks towards PSMA PET/CT imaging analysis tasks such as lesion classification, prediction of lymph node invasion, and segmentation.[21–23] However, current solutions are relatively nascent with limitations such as incorporating only imaging features and lack of clinical validation, and limited evidence for patient outcomes prediction. Deep learning methods incorporating multi-modal imaging and clinical data remains an active area of investigation.

Here we sought to develop a multi-modal deep learning framework that predicts PSA progression free survival (PFS) for mPCa patients from PSMA PET/CT imaging combined with clinical features. We present an approach that uses cropped images centered around detected lesions, focusing on tumor regions to minimize spurious correlations. Our method was also designed to improve prognostic performance through incorporation of clinical features in addition to imaging findings.

## Methods

### Data

This study included 396 lesions from 99 patients diagnosed with mPCa from a single academic institution. PSMA PET images were acquired using Ga-68 Gozetotide (PSMA-11) radiotracers that selectively bind to PSMA-expressing cells, enabling precise detection of metastatic lesions.[24] CT scans provide anatomical context for lesion localization, while binary lesion masks delineate regions of interest for automated analysis.

PSA progression was defined as two successive elevations of PSA that required re-staging or therapy change. Survival time was measured from initiation of systemic therapy to either the first PSA progression event or last follow-up, serving as the outcome variable for model training. Median follow up was 22.4 months. Clinical variables were identified by retrospective chart review.

### Image Resampling and Normalization

To ensure spatial consistency across all imaging data, PSMA PET, CT, and mask images were resampled to a standardized isotropic resolution of 1×1×1 mm with a uniform voxel grid of 256×256×256. PSMA PET and CT images were resampled using linear interpolation, while masks were resampled using nearest-neighbor interpolation to preserve binary values. Following resampling, images were aligned using affine transformation to maintain anatomic consistency. After resampling, image normalization was applied to standardize intensity distributions. PSMA PET images were normalized using the 2nd and 98th percentile of voxel intensity values to mitigate the impact of extreme outliers, while CT images were normalized by subtracting the Hounsfield Unit (HU) value of air (−1000 HU) and dividing by 1000 to scale the intensities.[25,26]

### Lesion Identification and Preprocessing

Regions containing putative malignant lesions were delineated by applying connected component analysis on each patient’s mask. For each detected lesion, we computed its center of mass and its spatial dimensions (depth, height, and width). The imaging volumes (PSMA PET, CT, and mask) were then cropped around each lesion. The crop size was dynamically adjusted based on the size of each lesion to include a box defined as twice the lesion size, ensuring that the entire lesion and surrounding context were captured. Individual lesion crops were padded to match the maximum lesion size and then concatenated along the depth dimension. To ensure uniform spatial dimensions across all patients, the concatenated volumes were further padded along the depth axis to a global maximum depth computed across all patients.

### Data Augmentation

To enhance sample diversity and address class imbalance, data augmentation was applied.[27] The augmentation strategy included random affine transformations, flipping, and noise injection. Random affine transformations were used to simulate variations in patient positioning by applying rotations of up to 90° and translations of up to 10 voxels along each axis. To introduce additional variability, images were randomly flipped along all three spatial axes. Furthermore, Gaussian random noise with a mean of 0 and a standard deviation of 0.1 was independently added to the PSMA PET and CT images. Due to a class imbalance with a censored-to-uncensored ratio of approximately 4:1, uncensored patients underwent five times more augmentations than censored patients, ensuring adequate representation of uncensored samples during training.

### Preprocessing of Clinical Features

We conducted univariate analyses using CoxPH regression to quantitatively assess the association of each clinical variable with PSA PFS with clinically relevant variables and those with statistically significant association at a *P*-value < 0.05 being considered candidates for further analysis. Those candidate variables were subsequently incorporated into a multivariate LASSO CoxPH regression model, which simultaneously evaluated all predictors while applying a regularization penalty to shrink coefficients and exclude non-contributory variables, mitigating multicollinearity and overfitting. Categorical variables were converted into a numerical format using one-hot encoding to ensure appropriate representation in the analysis. All other variables were also normalized to a range of 0 to 1. The k-nearest neighbors (KNN) algorithm was employed for missing data imputation.

### Model Architecture

The proposed survival prediction model is a multi-modal deep learning framework based on ResNet architecture as shown in **Figure 1**. ResNet is a convolutional neural network (CNN) architecture known for its use of residual blocks with skip connections.[28] Those connections add the input of a layer directly to its output, enabling the network to learn residual functions rather than full transformations, which mitigates the vanishing gradient problem and allows training of very deep networks.

**Figure 1.**
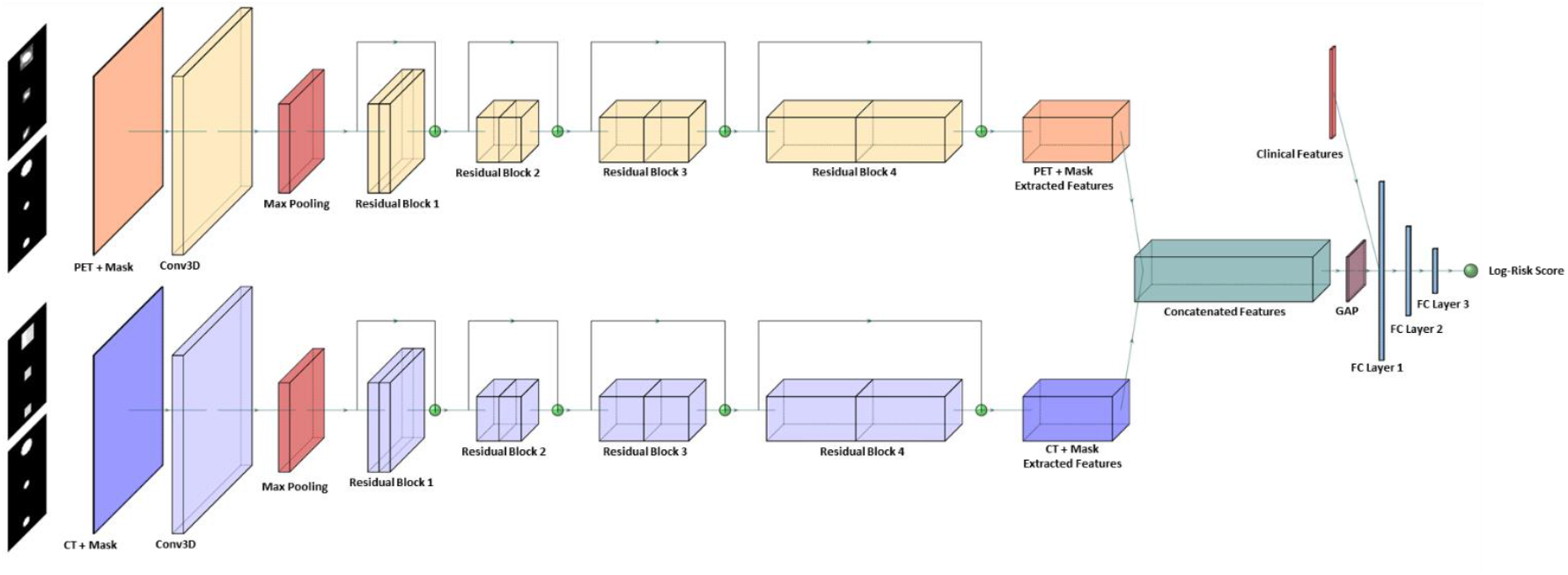
Architecture of the Multi-Modal Deep Learning Model for Survival Prediction using PSMA PET, CT, and Mask inputs

Our model uses a 3D ResNet backbone organized into four residual stages. It begins with a 7×7×7 convolutional layer with 64 filters, followed by batch normalization and 3D max-pooling to reduce spatial dimensions. In the initial stages, strided convolutions and pooling progressively downsample the feature maps. However, to preserve finer spatial details, the third and fourth stages employ dilated convolutions with dilation factors of 2 and 4, respectively. Each stage consists of multiple residual blocks built from 3×3×3 convolutional layers, batch normalization, and ReLU activations, with skip connections ensuring stable gradient flow.

Our model processes two imaging modalities, PSMA PET and CT, each accompanied by corresponding lesion bounding box masks. For each modality, the imaging data and its mask are concatenated along the channel dimension and passed through a dedicated instance of the 3D ResNet to extract modality-specific features. The feature vectors from both modalities are then concatenated to form a unified representation. A global average pooling layer is applied to condense the spatial information, and the resulting vector is combined with clinical features. Finally, the concatenated features pass through three fully connected layers—the first two use ReLU activations and the last outputs a single numerical log-risk score.

We employed the CoxPH loss function, which leverages a partial likelihood framework to estimate hazard ratios while inherently accounting for right-censored data [29]. This loss function assumes that the relative risk between any two patients stays constant over time. By optimizing the log-partial likelihood, the CoxPH loss ensures that patients who experience events earlier are assigned higher relative risk scores, thereby preserving the correct risk ranking.

### Training Procedure

Model training was conducted using a five-fold cross-validation strategy to ensure robust evaluation. To maintain the balance between censored and uncensored samples across folds, we ensured that each training fold contained 80% of the uncensored samples, while each validation fold retained the remaining 20% of uncensored patients. That stratified sampling strategy preserved the proportion of censored and uncensored cases across all folds, preventing potential biases in model evaluation. The models were trained from scratch using the Adam optimizer with batch size of 16, an initial learning rate of 10^-4^, a weight decay of 10^-4^, and a dropout rate of 0.3 in the fully connected layers. L1 regularization was applied with a weighting factor of 10^-4^ to encourage sparsity in learned features. A dynamic learning rate scheduler was applied to reduce the learning rate when the validation loss plateaued. All experiments were conducted on an NVIDIA A100 GPU.

### Evaluation Metrics and Statistical Analysis

The concordance index (C-index) was computed to measure how well the predicted risk scores corresponded to observed PSA PFS. A higher C-index indicates better agreement between predicted rankings and actual PSA PFS. To further assess the model’s clinical applicability, Kaplan Meier (KM) survival analysis was performed. Patients were stratified into high-and low-risk groups based on their predicted risk scores, and KM curves were generated to visualize differences in survival distributions. Log-rank tests were conducted to determine the statistical significance of survival differences between the predicted risk groups.

### Ethics Declarations

This retrospective study was approved by the UT Southwestern Medical Center Institutional Review Board. The Board waived the requirement for written informed consent because all data were de-identified prior to analysis. All methods were carried out in accordance with relevant guidelines and regulations, including the Declaration of Helsinki and the U.S. Common Rule. The PET/CT images shown are fully anonymized and contain no personally identifiable information.

## Results

### Baseline Characteristics of the Study Cohort

The cohort had a mean age of 68.5 ± 7.3 years. The majority of patients were Caucasian (78.7%), followed by African American (14.9%), Hispanic/Latino (3.2%), Asian (1.1%), and other races (2.1%). Most patients were non-smokers (81.7%), with a smaller proportion being former smokers (16.1%) or current smokers (2.2%). The median PSA nadir after treatment was 0.05 ng/mL (IQR: 0.05–0.27). At relapse, imaging, or initial staging, the median PSA level was 4.9 ng/mL (IQR: 0.43–13.84). Gleason scores were predominantly high, with 46.9% of patients having a Gleason score of 9, reflecting the aggressive nature of the disease in this cohort. Disease characteristics revealed that most patients had advanced clinical tumor stage (cT), with 52% classified as T3, and clinical nodal involvement (cN) was present in 34.7% of patients. The mean number of lesions per patient was 4 ± 2.6, with PSMA-positive lesions most commonly located in lymph nodes (51%). Notably, 11.1% of patients were castration-resistant at the time of imaging, and visceral lesions were present in 7.1% of patients. In terms of therapeutic interventions, 64.6% of patients underwent androgen deprivation therapy intensification (ADTi), and docetaxel was used in 5.1% of patients. Outcome data showed that 21.2% of patients experienced PSA progression, with a median PSA PFS of 22.4 months (**Table 1**).

**Table 1.** Baseline Characteristics of the Patients Included in the Study.

| Characteristic | Value or Distribution |
| --- | --- |
| Total Number of Patients | 99 |
| **Demographics** |  |
| Age, mean ± SD | 68.5 ± 7.3 |
| Race |  |
| Caucasian, n (%) | 74 (78.7%) |
| Hispanic/Latino, n (%) | 3 (3.2%) |
| African American, n (%) | 14 (14.9%) |
| Asian, n (%) | 1 (1.1%) |
| Other, n (%) | 2 (2.1%) |
| Smoking Status, n (%) |  |
| Non-smoker | 61 (81.7%) |
| Former smoker | 33 (16.1%) |
| Current smoker | 5 (2.2%) |
| BMI, mean ± SD | 28.1 ± 4.4 |
| **Clinical Characteristics** |  |
| ECOG Performance Status, n (%) |  |
| ECOG 0 | 76 (81.7%) |
| ECOG 1 | 15 (16.1%) |
| ECOG 2 | 2 (2.2%) |
| PSA Nadir (ng/mL) after Treatment, median [IQR] | 0.05 [0.05 - 0.27] |
| PSA at Relapse/Imaging/Initial Staging, median [IQR] | 4.9 [0.43–13.84] |
| Gleason Score, n (%) |  |
| Gleason Score 6 or 7 | 38 (38.8%) |
| Gleason Score 8 | 14 (14.3%) |
| Gleason Score 9 | 46 (46.9%) |
| **Disease Characteristics** |  |
| Clinical Tumor Stage, n (%) |  |
| T1 | 17 (22.7%) |
| T2 | 15 (20%) |
| T3 | 39 (52%) |
| T4 | 4 (5.3%) |
| Clinical Nodal Stage, n (%) |  |
| N0 | 49 (65.3%) |
| N1 | 26 (34.7%) |
| Number of Lesions, mean ± SD | 4 ± 2.6 |
| Castration Resistant, n (%) | 11 (11.1%) |
| PSMA-positive Lesion Location, n (%) |  |
| Lymph Node | 50 (51%) |
| Bone | 19 (19.4%) |
| Lymph Node and Bone | 29 (29.6%) |
| Visceral Lesions, n (%) | 7 (7.1%) |
| **Therapeutic Interventions** |  |
| No prior ADT | 84 (84.8%) |
| ADT after Diagnosis with PSMA PET | 99 (100%) |
| Hormonal Intensification, n (%) | 64 (64.6%) |
| Docetaxel Use, n (%) | 5 (5.1%) |
| **Outcomes** |  |
| PSA Progression, n (%) | 21 (21.2%) |
| PSA PFS, median | 22.4 |

### Clinical Feature Analysis

The univariate and multivariate LASSO CoxPH regression analyses showed that 10 clinical variables were statistically significant predictors of PSA PFS. Two additional variables—cN and ADTi—were retained based on their established clinical relevance in survival prediction. In addition, we included the binary variable castration resistance to differentiate hormone-sensitive from castration-resistant patients. Variables that did not meet these dual criteria were excluded from further consideration. Consequently, the final set of 12 clinical features incorporated into our deep learning prediction model comprised BMI, ECOG performance status, PSA nadir after treatment, PSA at relapse/imaging/initial staging, cT, cN, Gleason score, castration resistance, age, PSMA lesion location, ADT naïve, and ADTi.

### Comparison of Clinical and Imaging-Based Models

To assess the predictive performance of imaging data and clinical features in PSA PFS prediction, we compared three models: (1) a CoxPH model based only on clinical features, (2) an image-based deep learning model with ResNet-18 backbone without clinical features, and (3) an image-based deep learning model with ResNet-18 backbone that incorporates clinical features. Model performance was evaluated using the C-index across five-fold cross-validation, as summarized in **Table 2**. (ResNet backbone comparison shown in eSupplement Table 1.)

**Table 2.** C-index Performance Comparison of Clinical Features Only, Image-Based, and Hybrid Models Across Five-Fold Cross-Validation.

| Fold | Cox Model Based on Clinical Features | Image-Based Model without Clinical Features | Image-Based Model with Clinical Features |
| --- | --- | --- | --- |
| Fold 1 | 0.55 | 0.73 | 0.82 |
| Fold 2 | 0.57 | 0.72 | 0.74 |
| Fold 3 | 0.63 | 0.85 | 0.87 |
| Fold 4 | 0.60 | 0.70 | 0.78 |
| Fold 5 | 0.66 | 0.89 | 0.92 |

The CoxPH model using only clinical features demonstrated the lowest predictive performance across all five folds, with C-index values ranging from 0.55 to 0.66. That result indicated that while clinical variables provided prognostic information, their predictive capacity is limited when used alone. In contrast, the image-based deep learning model without clinical features achieved substantially higher C-index values, ranging from 0.70 to 0.89 across folds. This suggests that imaging-derived features capture critical prognostic information that is not fully reflected in traditional clinical features. The image-based deep learning model incorporating both imaging and clinical features consistently outperformed both the clinical-only and image-only models, achieving the highest C-index scores in every fold (0.74 to 0.92). This demonstrates the added predictive value of integrating clinical data with imaging features.

### Kaplan-Meier Survival Analysis and Risk Stratification

The KM curves stratified patients into low- and high-risk groups based on the median risk score cutoff, along with the min-max range (shaded regions in the plot) of survival probabilities across all folds, are presented in **Figure 2**. The median PSA PFS among the high-risk group was 19.7 months, compared to 26 months in the low-risk group (*P* < 0.001).

**Figure 2.**
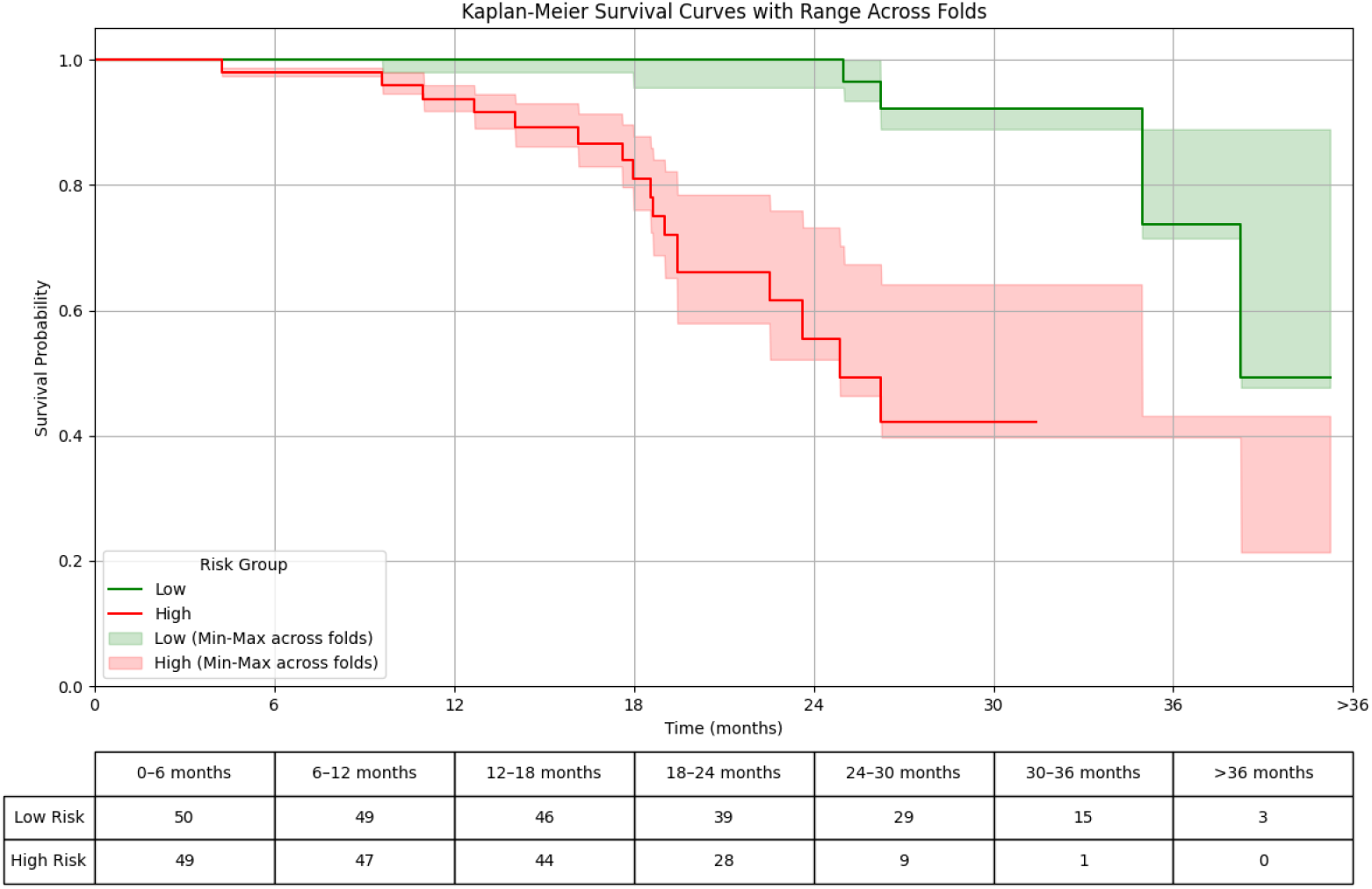
Kaplan Meier Survival Curves and At-Risk Table with Min-Max Range Across Folds: Low vs. High-Risk Group

To further examine the clinical relevance of the model’s predictions, PSMA PET/CT images of two representative patients, one from each risk category, are shown in **Figure 3**. The images display coronal, sagittal, and axial anatomical planes, with red bounding boxes highlighting detected lesions. The high-risk patient exhibits a greater number and larger lesions across multiple anatomical regions, including the pelvic lymph nodes, bones, and visceral organs with increased SUV uptake. In contrast, the low-risk patient presents with fewer and smaller metastatic lesions with relatively lower SUV values.

**Figure 3.**
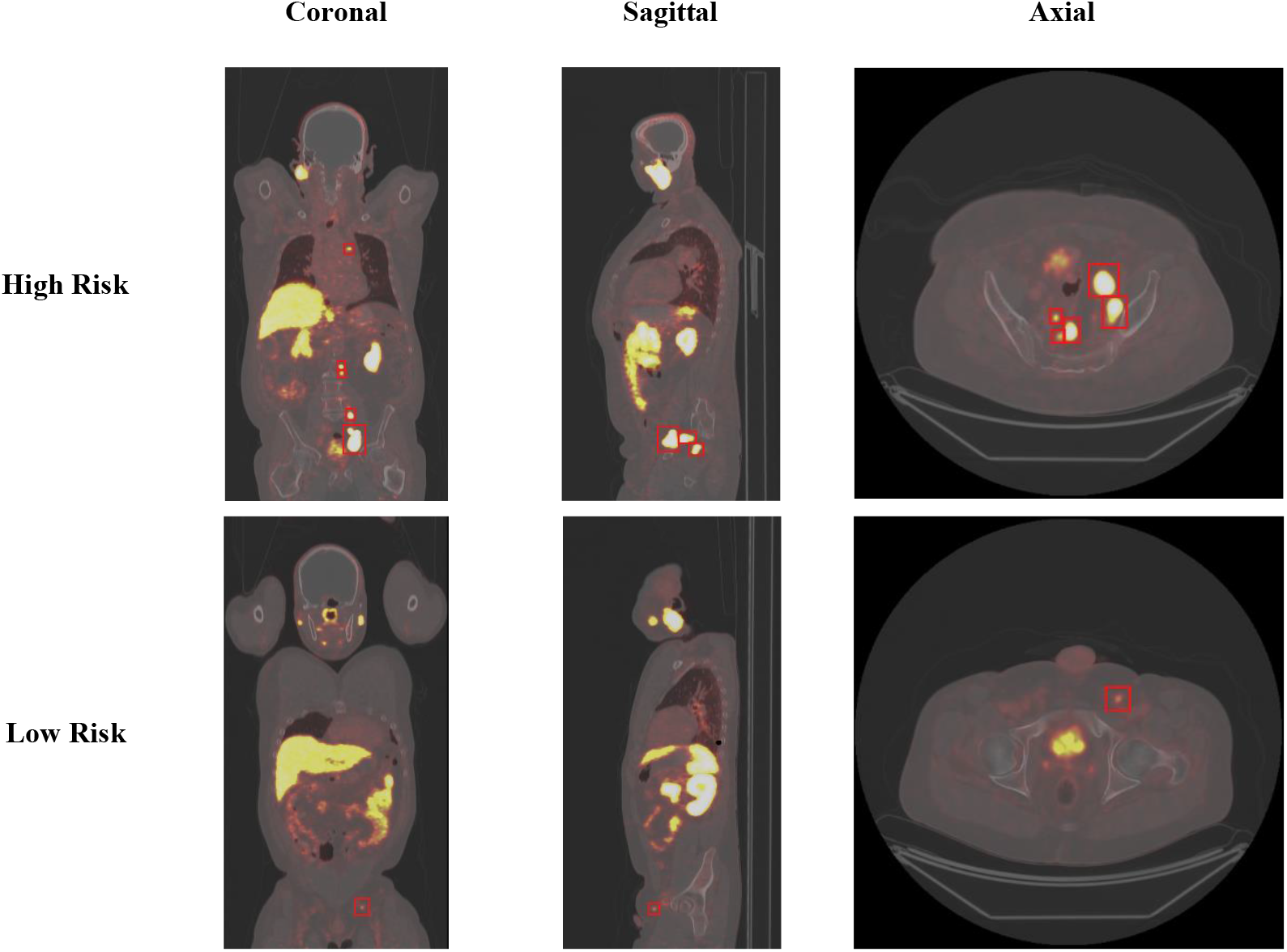
Representative Whole-Body PSMA PET/CT Images with Lesion Annotations for High-Risk and Low-Risk Patients

## Discussion

Our study demonstrates that a multi-modal deep learning framework integrating lesion-focused PSMA PET/CT imaging with key clinical features can effectively predict PSA PFS in patients with mPCa. The presented model, built on a ResNet-18 backbone, achieved high predictive accuracy across five-fold cross-validation and successfully stratified patients into high- and low-risk groups with significant differences in survival outcomes.

By integrating feature extraction and outcome prediction in a single pipeline, our deep learning framework can potentially overcome limitations of previous approaches that used handcrafted radiomic features followed by traditional machine learning models. The use of cropped whole-body PSMA PET/CT images centered on detected lesions reduces the possibility of spurious correlations by focusing on tumor-specific regions and eliminating extraneous background data. Incorporating clinical features selected via regression analyses further improved predictive accuracy, demonstrating that multimodal data fusion offers a more comprehensive representation of disease characteristics than either imaging or clinical data alone. This framework improves prognostication from PSMA PET/CT imaging and holds significant clinical implications, as accurate risk stratification may guide personalized treatment planning strategies for metastatic prostate cancer patients. For instance, oligometastatic patients with more favorable prognosis may benefit the most from local consolidation using ablative radiotherapy targeted to the primary tumor and metastatic sites and safely defer systemic therapy escalation.[30]

Our findings are corroborated by other studies that have demonstrated the potential for using AI to predict patient outcomes from PSMA imaging. Leung et al.[23] combined U-Net architectures for lesion segmentation and convolutional neural networks for feature extraction from cropped PET slices. The extracted features were then used to classify lesions into PSMA-RADS categories and to differentiate between benign and malignant lesions. In another study, Ma et al. [22] adopted a multimodal strategy by extracting features from [^68^Ga]Ga-PSMA-617 PET/CT images using the Med3D network, which were then combined with clinical features and standardized uptake value (SUV) measurements in a multi-kernel support vector machine to predict lymph node invasion. Seifert et al.[17] developed a prognostic risk score for overall survival prediction using PSMA PET-derived organ-specific tumor volumes. They employed a neural network for automated tumor segmentation, and the extracted tumor volumes underwent data preprocessing to derive radiomic features, which were then incorporated into Cox regression model for outcome prediction.

Despite promising results, our study has several limitations. We used retrospective data from a single-institution. The dataset is relatively limited in sample size, lacks detailed local therapy information, and has a short follow-up period reflected in the low event rate. Scanner and radiotracer-specific information was not well captured, so our models are unlikely to account for acquisition biases introduced by potential differences in equipment or imaging protocols. Furthermore, a heterogeneous cohort of mPCa patients was included in our analysis, and it is likely that further refinement of AI models for specific clinical scenarios such as de novo metastatic disease, metastatic recurrence, hormone-sensitive and castration-resistant settings will offer improved patient-specific predictions. Future research involving larger or prospective cohorts is necessary to further validate the clinical applicability of our framework.

In conclusion, we present a deep learning-derived imaging biomarker based on PSMA PET imaging for mPCa patients that demonstrates potential for prediction of PSA PFS prediction. By effectively integrating PSMA PET/CT imaging and clinical data while streamlining the feature extraction process, this approach has the potential to support more personalized treatment planning and improve patient outcomes.

## Data Availability

The data are not publicly available due to privacy or ethical restrictions. The data underlying this article may be made available on request from the corresponding author.

