## Supplemental Table 1 for "A multi-modal deep learning framework for predicting PSA progression-free survival in metastatic prostate cancer using PSMA PET/CT imaging"

**Table 1**. C-Index Performance Comparison of Survival Prediction Models with Different ResNet Backbones Across Five-Fold Cross-Validation

|  | ResNet-10-based Model | ResNet-18-based Model | ResNet-34-based Model |
| --- | --- | --- | --- |
| Fold 1 | 0.79 | 0.82 | 0.80 |
| Fold 2 | 0.71 | 0.74 | 0.72 |
| Fold 3 | 0.81 | 0.87 | 0.85 |
| Fold 4 | 0.77 | 0.78 | 0.78 |
| Fold 5 | 0.84 | 0.92 | 0.88 |
